# Dementia is strongly associated with medial temporal atrophy even after accounting for neuropathologies

**DOI:** 10.1101/2021.06.06.21258383

**Authors:** Davis C. Woodworth, Nasim Sheikh-Bahaei, Kiana A. Scambray, Michael J. Phelan, Mari Perez-Rosendahl, María M. Corrada, Claudia H. Kawas, S. Ahmad Sajjadi, for the Alzheimer’s Disease Neuroimaging Initiative

## Abstract

**Objective:** Brain atrophy is associated with degenerative neuropathologies as well as clinical status of dementia. Whether dementia influences atrophy independent of neuropathologies is not known. In this study, we examined the pattern of atrophy associated with dementia while accounting for the most common dementia-related neuropathologies.

**Methods:** We used data from National Alzheimer’s Coordinating Center (NACC, N=129) and Alzheimer’s Disease Neuroimaging Initiative (ADNI, N=47) participants with suitable in-vivo 3D-T1w MRI and autopsy data. We determined dementia status at visit closest to MRI. We examined the following dichotomized neuropathological variables: Alzheimer’s disease neuropathology, hippocampal sclerosis, Lewy Bodies, cerebral amyloid angiopathy, atherosclerosis. Voxel-based morphometry (VBM) identified areas associated with dementia after accounting for neuropathologies. Identified regions of interest were further analyzed. We used multiple linear regression models adjusted for neuropathologies and demographic variables.

**Results:** We found strong associations for dementia with volumes of the hippocampus, amygdala, and parahippocampus (semi-partial correlations≥0.28, P<0.0001 for all regions in NACC; semi-partial correlations≥0.35, P≤0.01 for hippocampus and parahippocampus in ADNI). Dementia status accounted for more unique variance in atrophy in these structures (∼8%) compared with neuropathological variables; the only exception was hippocampal sclerosis which accounted for more variance in hippocampal atrophy (10%).

**Conclusion:** Even after accounting for the most common neuropathologies, dementia still had among the strongest correlations with atrophy of medial temporal lobe structures. This suggests that atrophy of the medial temporal lobe is most related to clinical status of dementia as opposed to Alzheimer’s or other neuropathologies.

## INTRODUCTION

Some of the earliest imaging biomarkers proposed for Alzheimer’s disease (AD) were measures of brain atrophy. In 1988, Seab *et al* reported a 40% reduction in hippocampal volume in AD patients compared to control participants^1^. In 1989, De Leon *et al* found that in participants with symptoms of dementia, hippocampal atrophy, as assessed on computed tomography (CT) images, was more prevalent^2^. Since then, many studies have found associations between atrophy of brain regions and clinical AD, including some recurring regions such as structures of the medial temporal, lateral temporal, and parietal lobes, and these atrophy measures have been used for both diagnosis^3^ and as research biomarkers^4^ for AD.

Previous studies have also examined the association between brain atrophy, as assessed on imaging, and various neurodegenerative pathologies. An early study found that MRI- and autopsy-measured atrophy of the hippocampus were highly correlated and associated with lower hippocampal neuron counts^5^. Another early imaging-pathology correlation study, using participants from the Nun Study, found that hippocampal atrophy was significantly associated with Braak staging of neurofibrillary tangles^6^. Additional studies have found associations between AD neuropathology (ADNP) and whole brain as well as hippocampal atrophy, while other neuropathologies, such as hippocampal sclerosis of aging (HS) and TAR DNA-binding protein 43 (TDP-43), have also been found to be associated with hippocampal volumes and medial temporal lobe atrophy^7^.

Only a few studies have examined the association between atrophy on neuroimaging and clinical dementia while accounting for neuropathologies. The objective of this study was to examine the association between MRI-measured antemortem gray matter atrophy and clinical status of dementia while accounting for commonly assessed neuropathologies found at autopsy. To address this question we used data from participants with appropriate MRI and neuropathology data from the National Alzheimer’s Coordinating Center (NACC) as an exploration dataset, and the Alzheimer’s Disease Neuroimaging Initiative (ADNI) database as a validation dataset.

## METHODS

### Exploration dataset, NACC

We used data from participants with and without dementia, who had both MRI and pathology data available in the NACC database between September 2005 and March 2017. The NACC Uniform Data Set (UDS) consists of data submitted by approximately 30 National Institute on Aging (NIA) funded Alzheimer’s Disease Research Centers (ADRCs) across the USA^8^. Contributing ADRCs are approved by their local IRB. We identified 243 participants with both pathology data and at least one available MRI, and used 129 for our analyses (see *MRI* methods section below).

### Demographic and Clinical Variables, NACC Data

We selected dementia status, assigned by ADRC clinicians as either dementia of the Alzheimer’s type or other non-Alzheimer’s dementia, at the assessment closest to MRI as our variable of interest. We also selected the following demographic variables: sex, age at MRI, years from MRI to death, and years of education. We used Mini-Mental State Examination (MMSE) and Clinical Dementia Rating Sum of Boxes (CDR-SB) instead of dementia status for some analyses.

### Pathology

Neuropathological data are collected at the ADRCs via a standardized Neuropathology Form and Coding Guidebook^9, 10^. The NACC pathology data dictionary was used to define pathological categories. We assessed Alzheimer’s Disease neuropathology (ADNP), which we defined as present (+) based on high likelihood per the NIA-Reagan criteria^11^, comprising a combined Consortium to Establish a Registry for Alzheimer’s Disease (CERAD) neuritic plaque score of 3 (Frequent) and high Braak stage for neurofibrillary tangles (V/VI). We assessed hippocampal sclerosis (HS), defined as present/absent (+/-) based on either NPHIPSCL (NACC pathology form version 10), categorizing HS as present or absent (+/-) in the CA1 and/or subiculum, and NPSCL (NACC pathology form version 9 and earlier), categorizing medial temporal lobe sclerosis as present or absent (+/-) including hippocampal sclerosis. We also assessed: cerebral amyloid angiopathy (CAA) defined as none/mild (-) and moderate/severe (+); Lewy bodies, which we dichotomized as present/absent (+/-) based on presence in any region assessed; and atherosclerosis defined as none/mild (-) and moderate/severe (+). There was limited availability of hippocampal TDP-43 information for the participants in the data-freeze of NACC used for this study (23 of 129, or 18% of participants), and thus TDP-43 was not included for analyses. However, TDP-43 data was available for more ADNI participants and used for supplemental analysis (see *ADNI* section below). We also used the full range of severity (for atherosclerosis and CAA) and staging (for Braak tangles and CERAD plaques) scores as continuous variables for some analyses.

### MRI

**Supplementary Figure-1** shows a flow chart for inclusion and exclusion criteria of NACC participants. Out of the 243 participants with both pathology and at least one MRI available, 29 did not have a 3D-T1w scan. We made the decision to limit the data to scans that utilized an inversion-recovery (IR) preparation pulse, since this has a large effect on tissue contrast, especially between gray matter and white matter^12, 13^. There were no differences between participants with IR compared to non-IR sequences, which we report in the **Supplementary Methods**. Restricting the scans to those with IR sequences excluded another 74 participants rendering a sample of 140 participants with appropriate scans. To reduce time from scan to autopsy, for each participant we used the last available scan of sufficient quality.

The 3D-T1w IR scans were processed using the Computational Anatomy Toolbox (CAT12, http://www.neuro.uni-jena.de/cat/)^14^, which is implemented in the Statistical Parametric Mapping (SPM12, https://www.fil.ion.ucl.ac.uk/spm/software/spm12/) software. Preprocessing involved normalization, bias correction, and skull-stripping of the T1w images followed by tissue segmentation into gray matter (GM), white matter (WM), cerebrospinal fluid (CSF), and white matter hypointensities (WMH) based on the SPM12 tissue probability maps. Five participants failed segmentation: 1 participant was missing a substantial portion of brain tissue in the left lateral temporal lobe, and 4 participants had incorrect tissue class segmentation. Participants that failed processing were excluded from further analyses. An additional 6 participants were excluded due to missing data on one or more pathological variables. This left 129 participants as the final cohort used for analyses. For voxel-based morphometry (VBM) analyses, the GM density images were realigned to a common space, smoothed with a Gaussian filter with a full-width at half maximum of 8mm, and thresholded at a GM density of 0.05. For region of interest (ROI) analyses the unsmoothed but normalized GM density images were used, and volumes were extracted from ROIs defined by labels from the Harvard-Oxford cortical and subcortical atlases (maximum probability thresholds of 25%). Total intracranial volumes (TIV) were computed using CAT12. As a comparison to and validation for the CAT12 volume estimates, we also calculated hippocampal volumes using two other methods: FreeSurfer hippocampal subfield segmentation^15^, and Automatic Segmentation of Hippocampal Subfields (ASHS)^16^, which we outline in **Supplementary Methods**.

### Validation Dataset, ADNI

To replicate the results found in the NACC dataset, we used data from ADNI-1 participants who had both neuroimaging and pathology data available (N=47). ADNI (adni.loni.usc.edu) was launched in 2003, led by Principal Investigator Michael W. Weiner, MD, and has acquired serial MRI, positron emission tomography (PET), other biological markers, and clinical and neuropsychological assessment, to study the progression of mild cognitive impairment (MCI) and early Alzheimer’s disease (AD). For up-to-date information, see www.adni-info.org.

We selected ADNI-1 participants because the majority of participants with pathology data (47 out of 64 in the April 2018 public release) were ADNI-1. Additionally, the ADNI-1 MRI protocol consisted of an MPRAGE sequence on 1.5T scanners that renders the sequences more comparable across scanners. We used the clinical classification of AD according to ADNI criteria^17^ as our dementia variable. Briefly, physicians classified participants as AD based on clinical assessment, an MMSE score of less than 26, a clinical dementia rating (CDR) score of 0.5 or greater, and meeting NINCDS/ADRDA criteria^18^ for probable AD. We used the MRI closest to death for our analyses. Because ADNI neuropathology data are acquired following the NACC guidelines and forms (v10), we used the same neuropathological variables described in the *Pathology* Methods section above, with NPHIPSCL as the only variable for HS. For a supplemental analysis we used TDP-43 in the hippocampus (NPTDPC variable) which was available for a larger proportion of participants in ADNI (43 of 47, 91%) than NACC (18%).

### Statistics

To examine whether gray matter volume was associated with clinical status of dementia while accounting for neuropathologies, we performed a VBM analysis in the NACC dataset using multiple linear regression models on a voxel-wise basis, with dementia status, dichotomized neuropathological variables, sex, age at MRI, years of education, years from MRI to death, and TIV, as covariates. We used a voxel-wise threshold of P=0.001 and a cluster-size family-wise error rate threshold of P=0.05. Based on the voxel-wise VBM results, we then explored the relationship between the volumes of the various ROIs, as the dependent variables, with the demographic and neuropathological variables via multiple linear regression models. To verify the effects found using the dichotomized neuropathological variables, we performed a multiple linear regression in which we used severity scores for atherosclerosis and CAA as well as Braak stage and CERAD neuritic plaque scores as continuous variables. Also, to verify that the effects observed with dementia variable were present for more continuous measures of cognition, we performed multiple linear regressions using scores of MMSE and CDR-SB instead of dementia status. In order to further mitigate some of the scan-related heterogeneity, we examined multiple linear regressions in the subset of participants with magnetization-prepared rapid gradient echo (MPRAGE) scans (N=74), and those with a 3T MPRAGE scan (N=54), to observe whether the relative contributions of dementia, demographic, or neuropathological variables changed. For validation of the VBM-calculated hippocampal volumes, we performed Pearson’s correlations between these and those generated by FreeSurfer and ASHS, and also report semi-partial correlations for multiple linear regressions for hippocampal volumes generated by each of these techniques.

For replication we performed multiple linear regressions of the ROIs in the ADNI dataset. Since the publicly-available ADNI dataset is considerably smaller than the NACC, we used simplified statistical models by limiting the number of covariates to those found to be significantly associated (P<0.05) with the volumes of the ROIs in the NACC dataset. We compared the semi-partial correlation coefficients between the NACC and ADNI datasets as estimates of effect size and directionality. We next performed this same comparison except replacing the ADNP variable with Braak stage dichotomized as present for V/VI, as well as dichotomized by present for Braak stage of III/IV/V/VI. Lastly, we examined whether semi-partial correlation coefficients differed when using TDP-43 instead of HS, or when using both TDP-43 and HS in the same multiple linear regression.

We report the square of the semi-partial correlation as a measure of the unique variance in the dependent variable explained by an additional independent variable. It is the change in the multiple R-squared induced by the inclusion of an additional explanatory variable, after accounting for the contributions of all other independent variables in the model. This makes the semi-partial correlation and related variance explained more interpretable than partial correlations or standardized regression coefficients^19^.

### Data Availability

NACC and ADNI data are freely available to researchers upon request.

## RESULTS

### Participant Characteristics, NACC

The characteristics for the NACC participants are summarized in **Table-1**. There were 129 participants, most of whom presented with dementia at the time of MRI (94, 73%). Participants with dementia were significantly younger at time of MRI, were more likely to have ADNP, a higher Braak stage, higher CERAD neuritic plaque score, Lewy bodies, and more severe CAA. The 10^th^/90^th^ percentiles for age at MRI were 62years/85years for those with dementia and 70years/92years for those without dementia. There were no significant differences in terms of sex or years of education. Of note, around 75% of the participants without dementia had MCI at the time of MRI.

**Table-1.** Participant characteristics for NACC data. Continuous variables (age at MRI, years from MRI to death, and years of education) are reported as mean and standard deviations within each group and group differences are assessed by Student’s T-test. Categorical variables (sex, MCI, and pathological variables) are reported as number and percentage within each group and group differences are assessed by Chi-Squared tests. †Five participants with dementia were missing MMSE score. MCI: Mild cognitive impairment. MMSE: Mini-Mental State Examination. CDR-SB: Clinical Dementia Rating Sum of Boxes. ADNP: Alzheimer’s disease neuropathology. CERAD: Consortium to Establish a Registry for Alzheimer’s Disease. HS: Hippocampal Sclerosis. CAA: Cerebral amyloid angiopathy. Edu.: education.

| **Variable** | **Participants without Dementia (N=35)** | **Participants with Dementia (N=94)** | **Chi-square or T-test P-Value** |
| --- | --- | --- | --- |
| *Males* | 23 (66%) | 66 (70%) | 0.6 |
| *Age at MRI* | 82 (±9) years | 75 (±9) years | <0.001 |
| *Years Edu.* | 16 (±3) years | 16 (±3) years | 0.8 |
| *MCI* | 26 (74%) |  |  |
| *MMSE* | 26.7 (±2.6) | 18.8 (±6.4)<sup>†</sup> | <0.001 |
| *CDR-SB* | 1.7 (±1.7) | 7.7 (±4.2) | <0.001 |
| *Years MRI to Death* | 4 (±2) years | 3 (±2) years | 0.08 |
| *ADNP* | 11 (31%) | 61 (65%) | <0.001 |
| *Braak Stage 0/I/II* | 4 (11%) | 2 (2%) | 0.02 |
| *Braak Stage III/IV* | 8 (23%) | 12 (13%) |  |
| *Braak Stage V/VI* | 23 (66%) | 80 (85%) |  |
| *CERAD plaque 0* | 2 (6%) | 3 (3%) | 0.02 |
| *CERAD plaque 1* | 2 (6%) | 1 (1%) |  |
| *CERAD plaque 2* | 18 (51%) | 27 (29%) |  |
| *CERAD plaque 3* | 13 (37%) | 63 (67%) |  |
| *HS* | 6 (17%) | 16 (15%) | 0.7 |
| *Lewy Bodies* | 10 (29%) | 49 (52%) | 0.049 |
| *CAA* | 11 (31%) | 64 (68%) | 0.02 |
| *Atherosclerosis* | 14 (40%) | 30 (32%) | 0.3 |

### VBM Voxel-wise results, NACC

We observed strong associations between gray matter density and dementia in the hippocampus, amygdala, and adjacent temporal cortex, even after accounting for other demographic and neuropathological variables, with stronger associations in the left hemisphere (left cluster size=27.3mL, right cluster size=12.3mL, cluster-size family-wise error rate P<0.001 for both; **Figure-1.A**). The bilateral Harvard-Oxford regions with the highest percentage overlap with the VBM results (thresholded at P=0.001) were the amygdala (81%), hippocampus (64%), anterior parahippocampus (41%), and posterior parahippocampus (31%), **Figure-1.B**). The only other regions with significant overlap were the temporal pole (30%) and temporal fusiform gyrus (12%). We selected the medial temporal regions of the hippocampus, amygdala, and parahippocampus (consisting of the anterior and posterior parahippocampal regions) as ROIs to be used for further analysis. For comparison, we display VBM results that did not include neuropathological variables (using demographic variables only) in **Supplementary Figure-2**, which shows a similar pattern but with larger significant clusters that extend beyond the medial temporal lobe to the adjacent inferior and lateral temporal, as well as orbitofrontal lobes. *ROI Analysis, NACC*: The volumes of the ROIs (hippocampus, amygdala, parahippocampus) all showed significant and strong negative associations with dementia status, and when examining the unique variance explained by each of the variables, we found that dementia status had the strongest association for all the regions, accounting for the most unique variance (∼ 8%, **Table-2**). The one exception to this was HS, which accounted for more variance of hippocampal volume (∼10%). Additionally, we found significant associations between the volumes of these ROIs and age (greater atrophy with age), as well as TIV (greater intracranial volume associated with greater ROI volume). With regards to neuropathologies, the only significant associations with dementia were negative correlations between all ROIs and HS (strongest in the hippocampus with unique variance explained at ∼10%, lower in the other regions at ∼3%), and a negative correlation of amygdala volume with ADNP (2.3% variance explained) and Lewy bodies (2.9% variance explained). Adding dementia status to the models that already included the other variables increased the adjusted R^2^ from 0.34 to 0.42 for hippocampus, 0.35 to 0.43 for amygdala, and 0.27 to 0.35 for parahippocampus, and variance inflation factors for the model did not exceed 2 for any of the variables. Results were similar when using continuous instead of dichotomized measures for neuropathologies (**Table-3**). Results were also similar when, instead of dementia, MMSE (hippocampus sr=0.21, P=0.004; amygdala sr=0.21, P=0.008; parahippocampus sr=0.29, P<0.001) and CDR-SB (hippocampus sr=-0.22, P=0.001; amygdala sr=-0.24, P<0.001; parahippocampus sr=-0.34, P<0.001) were used in the model. Although dementia status did appear to have a slightly stronger effect (hippocampus sr=-0.28, amygdala sr=-0.29, parahippocampus sr=-0.28); effects for other demographic and neuropathological variables were not substantially different when using CDR-SB or MMSE (data not shown). When limiting the participants used for analysis to those with more homogeneous scans (MPRAGE and 3T MPRAGE), the effect of dementia on atrophy tended to increase, the effect of HS on atrophy tended to increase as well, and the effect of ADNP decreased in all regions and was no longer significant for the amygdala (**Table-4**). As validation, the VBM-generated hippocampal volume showed a high correlation with FreeSurfer (r=0.913, P<0.001) and ASHS (r=0.903, P<0.001) hippocampal volumes, and the multiple linear regressions using each of the different measures yielded similar patterns confirming the observed results, though the effects of variables and adjusted R^2^ were slightly reduced for the ASHS-generated hippocampal volumes (**Supplementary Table-1**).

**Figure. 1.**
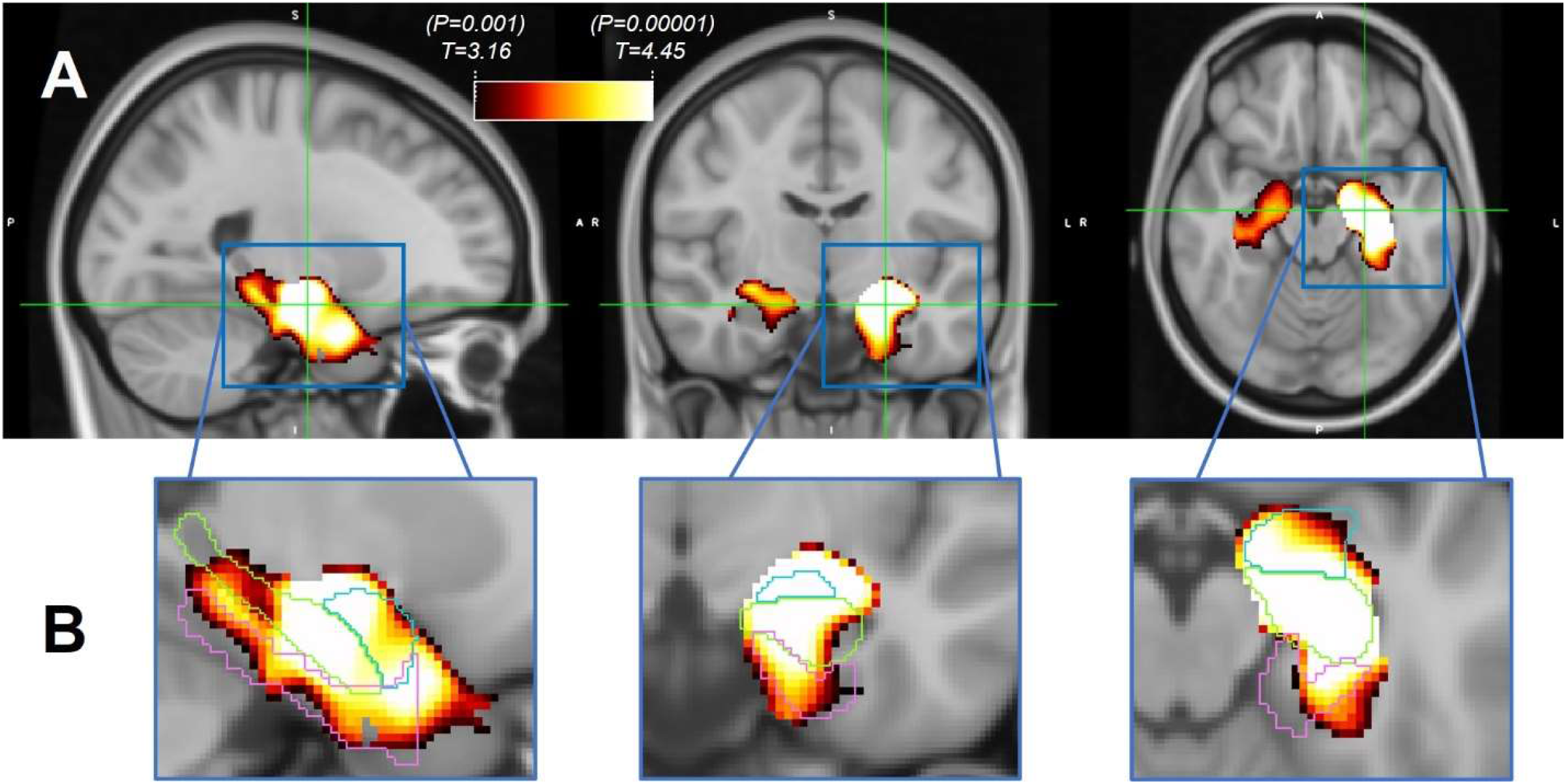
Voxel-wise VBM association gray matter atrophy with dementia in NACC data (N=129). Results thresholded at T-stat=3.16 (P=0.001) and survive cluster-wise false discovery rate threshold of 0.05. A) Strong associations present in the medial temporal lobe, with larger effects in the left hemisphere. B) Zoomed-in voxel-wise maps in left hemisphere with outlines of Harvard-Oxford atlas regions of the hippocampus (light green), amygdala (light blue), and parahippocampus (pink) illustrating degree of overlap between thresholded T-statistic maps and these regions. VBM: Voxel-based morphometry.

**Table-2.** Multiple linear regression results for ROI volumes across the various demographic and neuropathological measures for the NACC data (N=129). Bold denotes P<0.05. TIV: total intracranial volume. ADNP: Alzheimer’s disease neuropathology. HS: Hippocampal Sclerosis. CAA: Cerebral amyloid angiopathy. Semi-Part.: semi-partial correlation coefficient. Var. Exp.: unique variance explained.

| **Regions** |  | **Hippocampus** |  |  |  |
| --- | --- | --- | --- | --- | --- |
| **Variables** |  | **Semi-Part.** | **% Var. Exp.** | **T-stat** | **P-value** |
| **Demographic** | *Dementia Status at MRI* | **-0.28** | **7.78** | **-4.15** | **<0.001** |
|  | *Age* | **-0.27** | **7.51** | **-4.08** | **<0.001** |
|  | *Sex* | 0.05 | 0.20 | 0.67 | 0.5 |
|  | *Years Education* | 0.13 | 1.59 | 1.88 | 0.06 |
|  | *Years MRI to Death* | 0.11 | 1.25 | 1.67 | 0.1 |
|  | *TIV* | **0.19** | **3.61** | **2.84** | **0.005** |
| **Pathology** | *ADNP* | -0.06 | 0.41 | -0.96 | 0.3 |
|  | *HS* | **-0.31** | **9.73** | **-4.65** | **<0.001** |
|  | *Lewy Bodies* | -0.04 | 0.13 | -0.54 | 0.6 |
|  | *CAA* | -0.03 | 0.08 | -0.44 | 0.7 |
|  | *Atherosclerosis* | -0.07 | 0.44 | -0.98 | 0.3 |

| **Amygdala** |  |  |  |
| --- | --- | --- | --- |
| **Semi-Part.** | **% Var. Exp.** | **T-stat** | **P-value** |
| **-0.29** | **8.24** | **-4.30** | **<0.001** |
| **-0.24** | **5.95** | **-3.65** | **<0.001** |
| -0.04 | 0.18 | -0.64 | 0.5 |
| 0.12 | 1.42 | 1.78 | 0.08 |
| 0.09 | 0.88 | 1.41 | 0.2 |
| **0.19** | **3.50** | **2.80** | **0.006** |
| **-0.15** | **2.31** | **-2.27** | **0.03** |
| **-0.19** | **3.42** | **-2.77** | **0.006** |
| **-0.17** | **2.92** | **-2.56** | **0.01** |
| 0.12 | 1.46 | 1.81 | 0.07 |
| -0.08 | 0.67 | -1.22 | 0.2 |

| **Parahippocampus** |  |  |  |
| --- | --- | --- | --- |
| **Semi-Part.** | **% Var. Exp.** | **T-stat** | **P-value** |
| **-0.28** | **7.73** | **-3.91** | **<0.001** |
| **-0.25** | **6.30** | **-3.52** | **<0.001** |
| -0.12 | 1.35 | -1.62 | 0.1 |
| 0.04 | 0.12 | 0.49 | 0.6 |
| 0.11 | 1.10 | 1.48 | 0.1 |
| **0.15** | **2.28** | **2.13** | **0.04** |
| -0.06 | 0.35 | -0.83 | 0.4 |
| **-0.17** | **2.79** | **-2.35** | **0.02** |
| -0.09 | 0.79 | -1.25 | 0.2 |
| 0.05 | 0.23 | 0.68 | 0.5 |
| -0.08 | 0.59 | -1.08 | 0.3 |

**Table-3.** Multiple linear regression results for ROI volumes using full severity (CAA and atherosclerosis) or staging (Braak and CERAD) information as continuous variables in the NACC data (N=129). Bold denotes P<0.05. TIV: total intracranial volume. ADNP: Alzheimer’s disease neuropathology. HS: Hippocampal Sclerosis. CERAD: Consortium to Establish a Registry for Alzheimer’s Disease. CAA: Cerebral amyloid angiopathy. Semi-Part.: semi-partial correlation coefficient. Var. Exp.: unique variance explained.

| **Regions** |  | **Hippocampus** |  |  |  |
| --- | --- | --- | --- | --- | --- |
| **Variables** |  | **Semi-Part.** | **% Var. Exp.** | **T-stat** | **P-value** |
| **Demographic** | *Dementia Status at MRI* | **-0.29** | **8.18** | **-4.23** | **<0.001** |
|  | *Age* | **-0.32** | **10.50** | **-4.79** | **<0.001** |
|  | *Sex* | 0.03 | 0.10 | 0.46 | 0.6 |
|  | *Years Education* | 0.13 | 1.64 | 1.89 | 0.06 |
|  | *Years MRI to Death* | 0.09 | 0.76 | 1.28 | 0.2 |
|  | *TIV* | **0.18** | **3.10** | **2.60** | **0.01** |
| **Pathology** | *Braak Stage* | -0.05 | 0.23 | -0.71 | 0.5 |
|  | *CERAD Plaques* | 0.01 | 0.01 | 0.14 | 0.9 |
|  | *HS* | **-0.30** | **8.88** | **-4.41** | **<0.001** |
|  | *Lewy Bodies* | -0.04 | 0.14 | -0.55 | 0.6 |
|  | *CAA* | -0.05 | 0.22 | -0.70 | 0.5 |
|  | *Atherosclerosis* | -0.04 | 0.18 | -0.63 | 0.5 |

| **Amygdala** |  |  |  |
| --- | --- | --- | --- |
| **Semi-Part.** | **% Var. Exp.** | **T-stat** | **P-value** |
| **-0.30** | **8.88** | **-4.38** | **<0.001** |
| **-0.30** | **8.94** | **-4.40** | **<0.001** |
| -0.03 | 0.07 | -0.38 | 0.7 |
| 0.11 | 1.10 | 1.54 | 0.1 |
| 0.08 | 0.56 | 1.11 | 0.3 |
| **0.22** | **4.62** | **3.16** | **0.002** |
| 0.02 | 0.03 | 0.25 | 0.8 |
| **-0.14** | **1.88** | **-2.02** | **0.046** |
| **-0.20** | **3.92** | **-2.92** | **0.004** |
| **-0.15** | **2.25** | **-2.20** | **0.03** |
| 0.09 | 0.74 | 1.26 | 0.2 |
| -0.02 | 0.04 | -0.31 | 0.8 |

| **Parahippocampus** |  |  |  |
| --- | --- | --- | --- |
| **Semi-Part.** | **% Var. Exp.** | **T-stat** | **P-value** |
| **-0.27** | **7.08** | **-3.74** | **<0.001** |
| **-0.30** | **9.24** | **-4.28** | **<0.001** |
| -0.14 | 1.85 | -1.92 | 0.06 |
| 0.05 | 0.20 | 0.64 | 0.5 |
| 0.08 | 0.67 | 1.16 | 0.3 |
| 0.13 | 1.64 | 1.80 | 0.07 |
| -0.11 | 1.19 | -1.53 | 0.1 |
| 0.03 | 0.11 | 0.47 | 0.6 |
| **-0.15** | **2.22** | **-2.10** | **0.04** |
| -0.09 | 0.79 | -1.26 | 0.2 |
| 0.04 | 0.18 | 0.59 | 0.6 |
| -0.06 | 0.41 | -0.90 | 0.4 |

**Table-4.** Semi-partial correlation coefficients for ROI volumes using progressively more homogeneous scan subsets. Bold denotes P<0.05. TIV: total intracranial volume. ADNP: Alzheimer’s disease neuropathology. HS: Hippocampal Sclerosis. CAA: Cerebral amyloid angiopathy.

| Regions |  | Hippocampus |  |  | Amygdala |  |  | Parahippocampus |  |  |
| --- | --- | --- | --- | --- | --- | --- | --- | --- | --- | --- |
| Variables |  | All IR<br>(N=129) | MPRAGE<br>(N=74) | MPRAGE<br>3T<br>(N=54) | All IR<br>(N=129) | MPRAGE<br>(N=74) | MPRAGE<br>3T<br>(N=54) | All IR<br>(N=129) | MPRAGE<br>(N=74) | MPRAGE<br>3T<br>(N=54) |
| Demographic | Dementia Status at MRI | **-0.28** | **-0.39** | **-0.35** | **-0.29** | **-0.34** | **-0.30** | **-0.28** | **-0.35** | **-0.29** |
|  | Age | **-0.27** | **-0.23** | **-0.32** | **-0.24** | **-0.23** | **-0.26** | **-0.25** | **-0.24** | **-0.37** |
|  | Sex | 0.05 | -0.02 | -0.02 | -0.04 | -0.02 | 0.00 | -0.12 | -0.10 | -0.02 |
|  | Years Education | 0.13 | 0.17 | **0.22** | 0.12 | 0.09 | 0.11 | 0.04 | 0.09 | 0.12 |
|  | Years MRI to Death | 0.11 | 0.08 | 0.04 | 0.09 | 0.02 | -0.02 | 0.11 | -0.01 | -0.06 |
|  | TIV | **0.19** | **0.12** | 0.10 | **0.19** | **0.28** | **0.29** | **0.15** | **0.18** | **0.22** |
| Pathology | ADNP | -0.06 | -0.01 | -0.06 | **-0.15** | -0.05 | -0.06 | -0.06 | 0.04 | -0.02 |
|  | HS | **-0.31** | **-0.37** | **-0.40** | **-0.19** | **-0.23** | **-0.24** | **-0.17** | **-0.25** | **-0.26** |
|  | Lewy Bodies | -0.04 | 0.03 | 0.06 | **-0.17** | **-0.15** | -0.17 | -0.09 | -0.10 | -0.08 |
|  | CAA | -0.03 | -0.03 | 0.01 | 0.12 | 0.10 | 0.11 | 0.05 | 0.09 | 0.05 |
|  | Atherosclerosis | -0.07 | -0.10 | 0.01 | -0.08 | -0.07 | -0.01 | -0.08 | -0.10 | 0.03 |

### ROI Analysis, Comparison of NACC and ADNI

For the simplified statistical model to compare the NACC and ADNI data, we only used the following variables which were significant in the models using the NACC data: dementia, age, TIV, ADNP, HS, and Lewy bodies. **Table-5** displays the participant characteristics for the ADNI data, which were broadly similar those of the NACC dataset. Dementia was significantly associated with lower volumes of the hippocampus and parahippocampus, but not the amygdala where there was only a trend and relatively similar semi-partial correlations among dementia, HS, ADNP, and Lewy bodies (**Table-6**). The directionality of the findings for the neuropathological variables (ADNP and HS) were consistent for the ADNI and NACC datasets and broadly showed a similar magnitude, though ADNP had a larger (though not statistically significant) semi-partial correlation for each of the regions in the ADNI dataset. When using dichotomized Braak stage instead of ADNP, the trends were largely similar, with nearly identical findings when using Braak Stage V/VI or Braak stage III/IV/V/VI classifications (**Supplementary Table-2**). Lastly, the effect of dementia did not change when using TDP-43 instead of HS or when using both in the same model (**Supplementary Table-3**). TDP-43 tended to show similar effects when used instead of HS but the semi-partial correlations for TDP-43 and adjusted R^2^ for the model were lower, and when including both TDP-43 and HS in the same model HS tended to eclipse the effects of TDP-43.

**Table-5.**
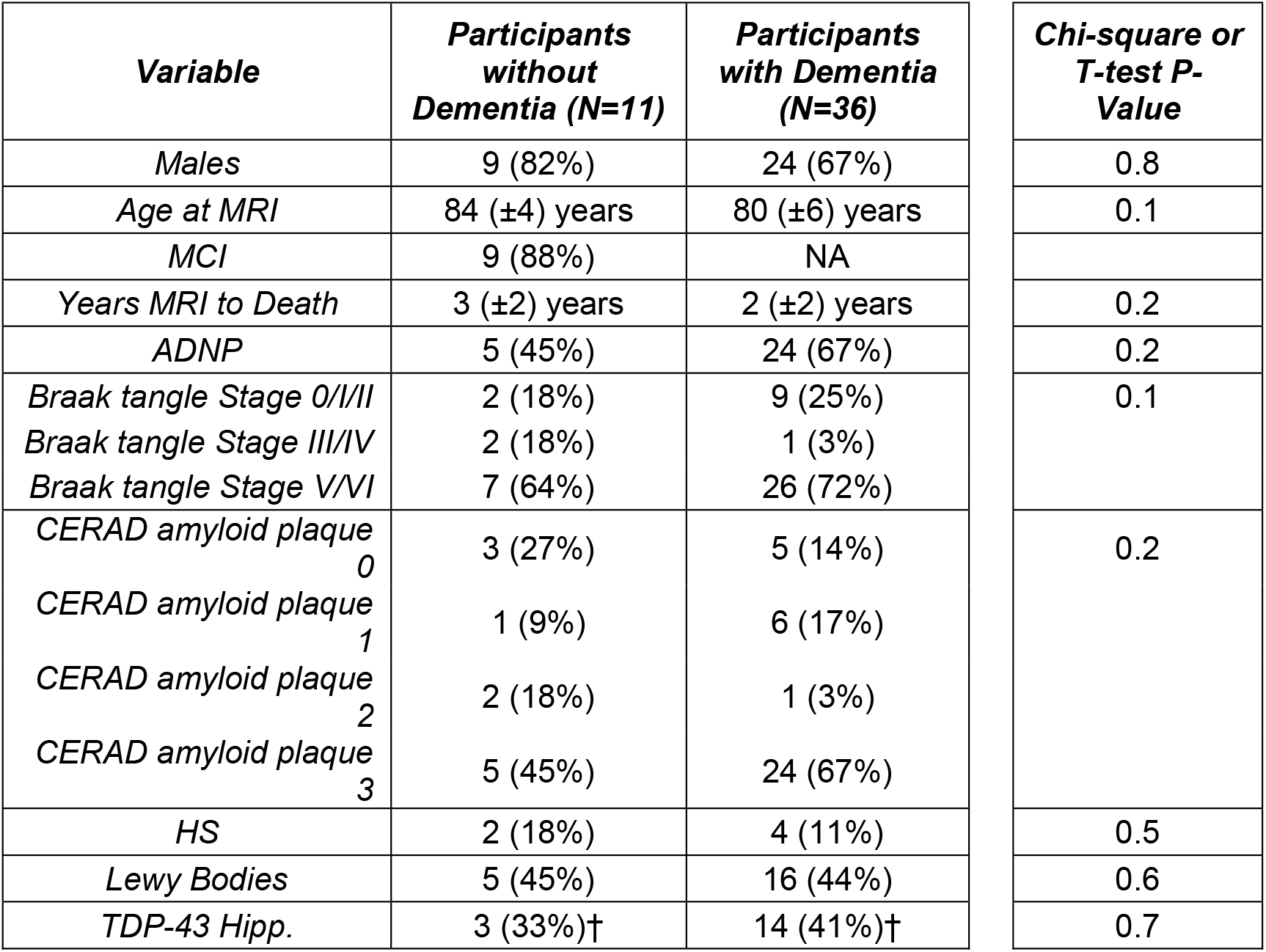
Participant characteristics of ADNI data. Continuous variables (age at MRI, years from MRI to death) are reported as mean and standard deviation within each group and group differences are assessed by Student’s T-test. Categorical variables (sex and pathological variables) are reported as number and percentage within each group and group differences are assessed by Chi-Squared tests. †Two participants with and two participants without dementia were missing TDP-43 information in the hippocampus. MCI: Mild Cognitive Impairment. ADNP: Alzheimer’s Disease Neuropathology. CERAD: Consortium to Establish a Registry for Alzheimer’s Disease. HS: Hippocampal Sclerosis. TDP-43: TAR DNA-binding protein 43. Hipp.: Hippocampus.

**Table-6.** Comparison of NACC (N=129) and ADNI (N=47) semi-partial correlation coefficients for ROI volumes across the various demographic and neuropathological measures. Bold denotes P<0.05. ROI: Region of interest. TIV: total intracranial volume. ADNP: Alzheimer’s disease neuropathology. HS: Hippocampal Sclerosis. Semi-Part.: semi-partial correlation coefficient.

| **Regions** | **Hippocampus**<br>**Semi-Part. Corr.** |  | **Amygdala**<br>**Semi-Part. Corr.** |  | **Parahippocampus**<br>**Semi-Part. Corr.** |  |
| --- | --- | --- | --- | --- | --- | --- |
| **Variables** | *NACC* | *ADNI* | *NACC* | *ADNI* | *NACC* | *ADNI* |
| *Dementia* | **-0.34** | **-0.35** | **-0.30** | -0.21 | **-0.31** | **-0.35** |
| *Age* | **-0.37** | 0.11 | **-0.34** | 0.12 | **-0.34** | 0.11 |
| *TIV* | **0.28** | 0.23 | **0.38** | 0.19 | **0.33** | 0.09 |
| *ADNP* | -0.05 | -0.18 | -0.15 | -0.21 | -0.05 | -0.21 |
| *HS* | **-0.28** | **-0.47** | **-0.18** | -0.28 | **-0.17** | **-0.35** |
| *Lewy Bodies* | -0.05 | **-0.29** | **-0.17** | -0.27 | -0.07 | -0.22 |

## DISCUSSION

We examined the relationship between gray matter atrophy and clinical dementia status while accounting for other demographic and commonly-assessed neuropathological variables using the NACC and ADNI databases. We found a strong association between dementia and atrophy of medial temporal lobe structures, namely the hippocampus, amygdala, and parahippocampus, even after accounting for neuropathologies. This association of medial temporal lobe atrophy with dementia was stronger than for any other demographic or neuropathological variables, except for hippocampal sclerosis in relation to hippocampal volume. We found similar direction and size for the effect of dementia on atrophy for: 1) models using both dichotomized and continuous measures for neuropathologies, 2) for both dementia status as well as more continuous measures of cognitive burden (MMSE and CDR-SB), 3) for hippocampal volumes generated from three different pipelines (CAT12, FreeSurfer, ASHS), 4) for subsets of participants with more homogeneous MRI acquisitions, and 5) across two different imaging-pathology databases (NACC and ADNI). The consistency in the results across these methodological variations underscores the strength and robustness of our findings. The clinical implication of this finding is that medial temporal lobe atrophy should not necessarily be construed as a surrogate marker for degenerative pathologies (with the potential exception of hippocampal sclerosis) but rather as a more general indicator of neuronal damage and loss that can be caused by a variety of known, and other potentially unknown, factors.

Although the observed association of the clinical status of dementia and medial temporal lobe atrophy is not surprising and previously reported by many other groups, we found that this effect remains after accounting for neuropathologies. Perhaps surprisingly, we found the effect of dementia on medial temporal lobe atrophy was greater than that of most of the neuropathologies. Some studies using in vivo biomarkers for Alzheimer’s pathology have found similar results, such as stronger associations between hippocampal volume and cognition compared to CSF amyloid^20^, neurodegeneration measured by FDG-18 PET being a stronger predictor of cognition than amyloid PET^21^, and atrophy attenuating the effects of CSF amyloid and tau on cognition^22^. These *in vivo* measures of Alzheimer’s disease offer important information during life. However, in our study we used neuropathological ratings assigned at autopsy, including pathologies other than Alzheimer’s, which are considered the gold standard.

While many studies have examined the relationship between brain atrophy and either cognitive status or neuropathological findings separately, far fewer have assessed these relationships in a contiguous fashion. Among studies that have assessed both aspects, there have been conflicting results. In a paper by the Nun Study on 56 participants, hippocampal volume from imaging was related to both cognitive status and ADNP, both appearing to have an additive effect on atrophy^6^. Another study examined the relationship between MRI-derived hippocampal volumes in a cohort of 15 participants without and 24 participants with dementia^23^. They found hippocampal volume closest to death was related to hippocampal neurofibrillary tangle load in participants with dementia, but not in those without, and when dementia duration was taken into consideration the relationship between neurofibrillary tangles and hippocampal volumes disappeared^23^. A later study on 36 participants, all with neurofibrillary tangle stage V/VI and CERAD amyloid plaque score 2/3 (12 with normal cognition and 24 with dementia), found that participants who did not present with dementia had significantly larger total brain and hippocampal volumes, even after adjusting for common demographic factors^24^. Another study using visual assessments of postmortem MRI scans of brains in the Vantaa 85+ study found that MTA was not specific for ADNP, but was specific for dementia diagnosis^25^. A Mayo clinic study in 67 participants found that hippocampal volume was correlated with Braak stage (r=-0.39), but was more strongly correlated with MMSE scores (r=0.60)^26^. In this same study hippocampal volume was only associated with Braak stage in the overall sample and in participants with clinical AD (median Braak stage of 5, range 2-6), while within normal aging participants there was no significant relationship (median Braak stage of 2, range 0-4). Another study found that while hippocampal volume was among the best discriminators of clinical status, it performed poorly in its association with Braak stage where instead the volume of the inferior temporal lobe had the strongest association^27^. Thus, while previous studies have found associations between ADNP and medial temporal lobe atrophy, at least some of these studies find mitigated relationships in comparison to, or in the context of, dementia.

In our analysis of the NACC data that included more participants than any previous studies, after accounting for dementia, the presence of ADNP was either weakly or not significantly associated with volumes of the medial temporal lobe structures. This result was true when using ADNP, Braak stage as a continuous or dichotomized variable, for different pipelines for segmenting the hippocampus, and across both NACC and ADNI datasets. Previous studies have suggested that neuronal death is a sufficient condition for an amnestic type syndrome typically associated with AD: a study by Ball *et al* published in 1985 found that in participants with clinical AD, neuronal loss and gliosis were the only consistent findings^28^. Our results seem to coincide with this train of thought, namely that neuronal death appears to be the cause of medial temporal lobe atrophy, but high burden of ADNP does not necessarily result in this neuronal death and atrophy if there is no clinical presentation of dementia. A recent a study using multiple linear regression models found that while ADNP explained 6% of hippocampal volume in the whole cohort, when split into participants with and participants without dementia the variance explained by ADNP dropped to 3% for each of these subgroups^29^, suggesting that the effect of ADNP on hippocampal atrophy was tightly linked to dementia. This study also found that the effect of HS and TDP-43 were more pronounced than that of ADNP and also found similar results when using dichotomized versus continuous versions of neuropathological variables, which are similar to the findings in our study.

A recent study examining hippocampal volumes on postmortem MRI using Religious Orders Study (ROS) and Memory and Aging Project (MAP) data found that, after accounting for demographic and neuropathological variables, the addition of hippocampal volumes to their models explained an additional 5% in the variance of cognitive decline in participants^30^. Our results are in agreement with the ROS/MAP study. Additionally, we believe our study contributes the following: 1) In our study, we examined the contribution of dementia and neuropathologies to gray matter atrophy on brain MRI. The ROS/MAP study examined the contribution of hippocampal volume on MRI to cognitive decline in an attempt to investigate the mechanisms underlying this decline. Our study arguably provides a more relevant clinical perspective from the standpoint of interpreting the common radiological finding of medial temporal atrophy. 2) The MRIs used for our study were *in vivo*, reminiscent to the information clinicians acquire to help them predict the underlying explanation for cognitive impairment, while the ROS/MAP study used postmortem MRI. 3) The MRIs used for our study spanned a wide variation in scanners and sequences, yet we still found strong associations that survived the potential noise introduced by the aforementioned heterogeneity. The ability to reproduce this finding with a more heterogeneous data confirms its utility in clinical setting.

Our results of associations between medial temporal lobe structures and hippocampal sclerosis of aging are also of interest. Many previous studies have noted that HS is associated with hippocampal atrophy^31-33^ and one study has even noted associations outside the medial temporal lobe^34^. Also, other studies have found TDP-43, the proteinopathy signature of HS, to be associated with hippocampal^29^ and additional brain atrophy^35^. However, it is unclear how the relationship between HS pathology and atrophy may change when accounting for cognitive status. We found a strong association of HS with hippocampal volume (accounting for ∼10% of variance), while also finding some associations with amygdala and parahippocampal volume, even when accounting for dementia. This finding underscores the relevance of hippocampal volume as a potential biomarker for HS. Additionally, our analyses in a subset of ADNI participants with TDP-43 information indicated that while TDP-43 behaves similar to HS, its effects are weaker. However, this analysis was in a smaller subset of participants and represents a limitation in this study: further research in larger datasets should examine medial temporal lobe atrophy in relation to cognition while accounting for more complete TDP-43 staging.

One of the limitations of this study is the heterogeneity of the MRI data, specifically in the NACC dataset. We did, however, limit the sequence paradigms used in the analysis to those that implemented an inversion recovery pulse, as it results in the greatest difference in tissue contrast. We also performed analyses limited to subsets of more homogeneous scans and the associations in these analyses tended to be slightly stronger and appeared to confirm the results in the full dataset. Additionally, the ADNI dataset circumvented some of these confounding factors by implementing a similar sequence across different scan vendors, all at 1.5T. Also, the similar strength of associations with cognitive status in both datasets (a more heterogeneous and a more homogeneous sample) highlights the robustness of our finding and the potential clinical application of the results. Another limitation to this study is the high proportion of participants with dementia and with high Braak stage and CERAD neuritic plaque scores in the NACC data freeze used. While the ADNI dataset partially circumvented this issue, further studies should examine these relationships in datasets with a more balanced proportion of participants without dementia and lower ADNP pathology burden. However, this is a limitation present in many neuropathology datasets where a large proportion of participants have dementia and a high ADNP burden. One final limitation of the study is one that is germane to most dementia research in that the participants used in this study are not necessarily a representative sample of the community, limiting generalizability.

In conclusion, we found a significant association between atrophy of medial temporal lobe structures, namely the hippocampus, amygdala, and parahippocampus, and clinical status of dementia, even while accounting for many of the commonly-assessed neuropathologies. This association of medial temporal lobe atrophy with dementia was stronger than any other demographic or neuropathological variable, with the exception of hippocampal sclerosis in relation to hippocampal volume. Alzheimer’s disease neuropathology was not associated with hippocampal volume but was weakly associated with amygdala volume in our models. These findings have significant implications on the diagnostic utility of MRI in dementia clinics and confirm the significant pathology-independent relationship between dementia and atrophy of structures that are generally considered to be the hallmark of Alzheimer’s disease.

## Data Availability

National Alzheimer’s Coordinating Center (NACC) and Alzheimer’s Disease Neuroimaging Initiative (ADNI) data are freely available to researchers upon request.

## Acknowledgments

The NACC database is funded by NIA/NIH Grant U01 AG016976. NACC data are contributed by the NIA-funded ADCs: P30 AG019610 (PI Eric Reiman, MD), P30 AG013846 (PI Neil Kowall, MD), P30 AG062428-01 (PI James Leverenz, MD) P50 AG008702 (PI Scott Small, MD), P50 AG025688 (PI Allan Levey, MD, PhD), P50 AG047266 (PI Todd Golde, MD, PhD), P30 AG010133 (PI Andrew Saykin, PsyD), P50 AG005146 (PI Marilyn Albert, PhD), P30 AG062421-01 (PI Bradley Hyman, MD, PhD), P30 AG062422-01 (PI Ronald Petersen, MD, PhD), P50 AG005138 (PI Mary Sano, PhD), P30 AG008051 (PI Thomas Wisniewski, MD), P30 AG013854 (PI Robert Vassar, PhD), P30 AG008017 (PI Jeffrey Kaye, MD), P30 AG010161 (PI David Bennett, MD), P50 AG047366 (PI Victor Henderson, MD, MS), P30 AG010129 (PI Charles DeCarli, MD), P50 AG016573 (PI Frank LaFerla, PhD), P30 AG062429-01(PI James Brewer, MD, PhD), P50 AG023501 (PI Bruce Miller, MD), P30 AG035982 (PI Russell Swerdlow, MD), P30 AG028383 (PI Linda Van Eldik, PhD), P30 AG053760 (PI Henry Paulson, MD, PhD), P30 AG010124 (PI John Trojanowski, MD, PhD), P50 AG005133 (PI Oscar Lopez, MD), P50 AG005142 (PI Helena Chui, MD), P30 AG012300 (PI Roger Rosenberg, MD), P30 AG049638 (PI Suzanne Craft, PhD), P50 AG005136 (PI Thomas Grabowski, MD), P30 AG062715-01 (PI Sanjay Asthana, MD, FRCP), P50 AG005681 (PI John Morris, MD), P50 AG047270 (PI Stephen Strittmatter, MD, PhD).

ADNI data collection and sharing for this project was funded by the Alzheimer’s Disease Neuroimaging Initiative (ADNI) (National Institutes of Health Grant U01 AG024904) and DOD ADNI (Department of Defense award number W81XWH-12-2-0012). ADNI is funded by the National Institute on Aging, the National Institute of Biomedical Imaging and Bioengineering, and through generous contributions from the following: AbbVie, Alzheimer’s Association; Alzheimer’s Drug Discovery Foundation; Araclon Biotech; BioClinica, Inc.; Biogen; Bristol-Myers Squibb Company; CereSpir, Inc.; Cogstate; Eisai Inc.; Elan Pharmaceuticals, Inc.; Eli Lilly and Company; EuroImmun; F. Hoffmann-La Roche Ltd and its affiliated company Genentech, Inc.; Fujirebio; GE Healthcare; IXICO Ltd.;Janssen Alzheimer Immunotherapy Research & Development, LLC.; Johnson & Johnson Pharmaceutical Research & Development LLC.; Lumosity; Lundbeck; Merck & Co., Inc.;Meso Scale Diagnostics, LLC.; NeuroRx Research; Neurotrack Technologies; Novartis Pharmaceuticals Corporation; Pfizer Inc.; Piramal Imaging; Servier; Takeda Pharmaceutical Company; and Transition Therapeutics. The Canadian Institutes of Health Research is providing funds to support ADNI clinical sites in Canada. Private sector contributions are facilitated by the Foundation for the National Institutes of Health (www.fnih.org). The grantee organization is the Northern California Institute for Research and Education, and the study is coordinated by the Alzheimer’s Therapeutic Research Institute at the University of Southern California. ADNI data are disseminated by the Laboratory for Neuro Imaging at the University of Southern California.

