## Supplementary figures and images for "Dementia is strongly associated with medial temporal atrophy even after accounting for neuropathologies"

### Supplemental Figure 1

**Supplementary-Figure-1.** *Flowchart of inclusion criteria for NACC participants*.

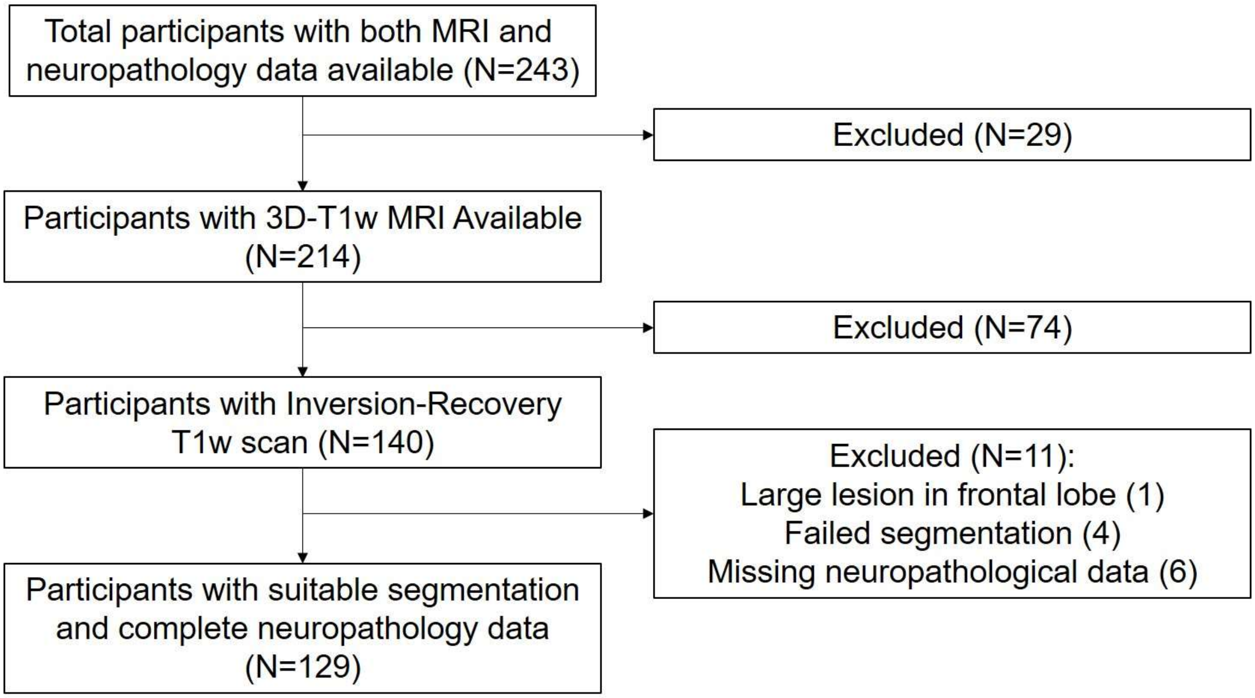
