## Supplemental Figure 2 for "Dementia is strongly associated with medial temporal atrophy even after accounting for neuropathologies"

**Supplementary-Figure-2.** *Voxelwise VBM association of dementia with gray matter atrophy accounting for demographic and neuropathology variables, accounting for demographic variables only, and outlines of thresholded maps, in NACC data (N=129)*. Results thresholded at T-stat=3.16 (P=0.001) and survive cluster-wise false discovery rate threshold of 0.05. A) Association between gray matter and dementia status accounting for demographic and neuropathology variables. B) Association between gray matter and dementia status accounting for demographic variables only. C) Outlines of both, showing more widespread significant regions for model accounting for demographic variables only.

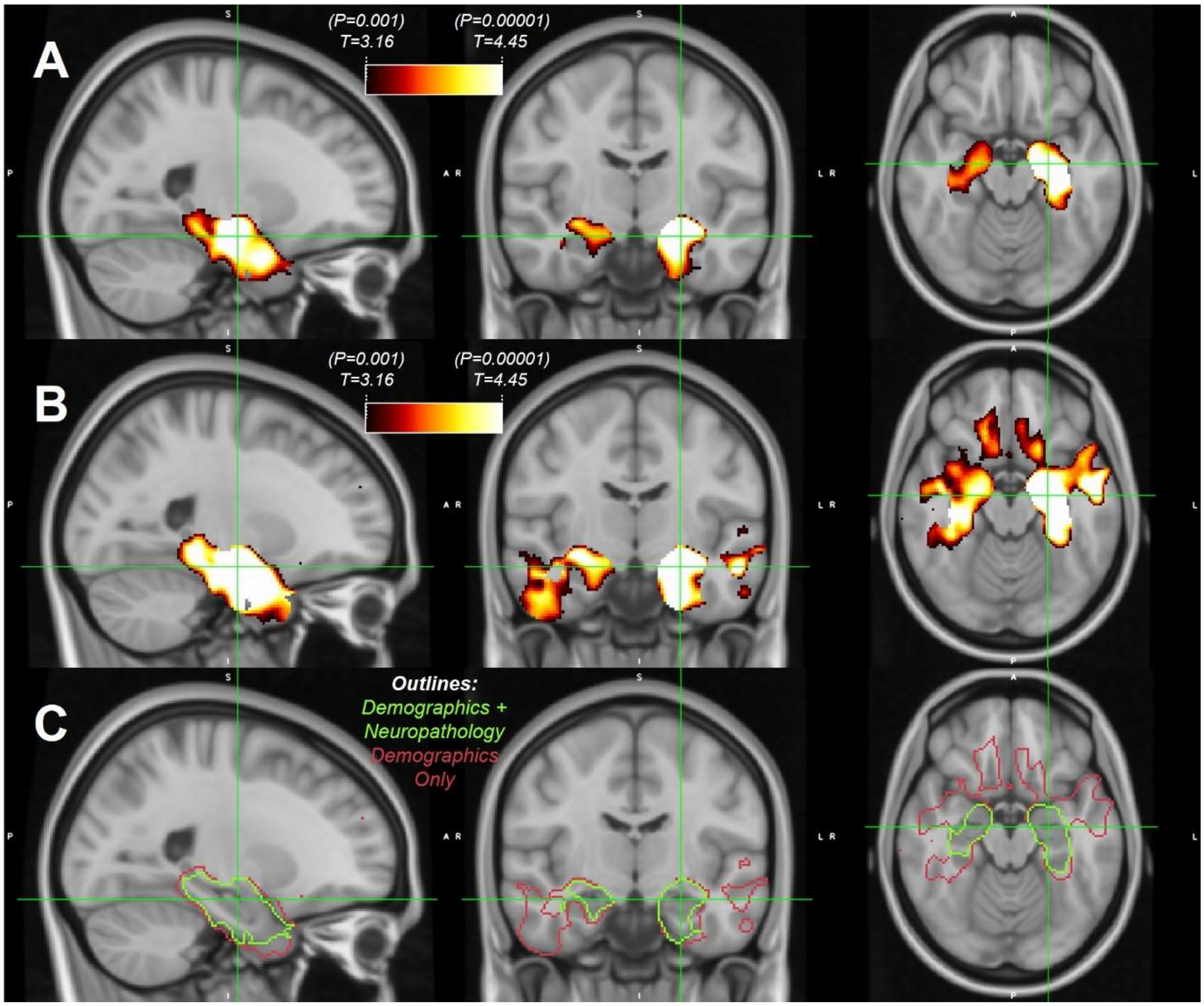
