## Supplemental Methods for "Dementia is strongly associated with medial temporal atrophy even after accounting for neuropathologies"

**Supplementary Methods.** *Information regarding selection of IR scans:* In National Alzheimer's Coordinating Center (NACC) database there is a wide variation in MRI scanner vendor, model, field strength, and acquisition parameters for 3D T1w scans. Inversion recovery (IR) sequences comprised inversion-recovery Fast Spoiled Gradient Recalled Echo (IR-FSPGR) on General Electric (GE) scanners and magnetization-prepared rapid acquisition gradient echo (MPRAGE) on GE or Siemens scanners. We selected these anticipating that they would present with improved gray/white matter contrast compared to non-IR pulse sequences. This was verified by visual assessment (data not shown) and the finding that, on average, non-IR scans presented with 7% less gray matter than the IR scans ($P<0.001$, Student's T-test). There were no differences between participants with and without IR scans in terms of number of participants with dementia ($P=0.7$), ADNP ($P=0.8$), HS ($p=0.7$, Chi-squared tests), or in terms of age at MRI ($P=0.4$) or years from MRI to death ($P=0.14$). Additionally, since ADNI data consists of IR scans only, limiting the NACC data to IR scans allowed for a better comparison between the datasets.

*Additional hippocampal segmentation pipelines:* In addition to the Computational Anatomy Toolbox (CAT12) voxel-based morphometry (VBM) estimates of volume for the hippocampus, the MRI scans for the same participants were processed using the FreeSurfer hippocampal subfield segmentations and the Automatic Segmentation of Hippocampal Subfields (ASHS) Penn Memory Center T1-Only Atlas for T1-weighted 3T MRI (PMC-T1) pipelines. All MRIs were 3D T1w scans with roughly $1mm^3$ isotropic resolution and full brain coverage, thus suitable for processing through both pipelines. Ultimately 120 out of the 129 total participants had suitable segmentations using all three pipelines and data from these 120 participants was used for correlation and multiple linear regression analyses comparing the volumes generated by the techniques.

*FreeSurfer Hippocampal Pipeline*: FreeSurfer v6.0 was used. First, standard FreeSurfer processing was performed through the *recon-all* command. Then, FreeSurfer hippocampal subfield segmentation module (https://surfer.nmr.mgh.harvard.edu/fswiki/HippocampalSubfields) was run. All segmentations were visually inspected for correctness of hippocampal estimates (i.e. exclusion of surrounding cerebrospinal fluid, white matter, and brainstem). For scans that did not process correctly due to *recon-all* failure, alternative skullstrip or Talairach registration steps were performed. Volume estimates came from the segmentation files and addition of the whole hippocampal volumes for the left and right hemispheres.

*ASHS PMC-T1 Pipeline*: ASHS produces segmentations of medial temporal lobe structures, including the anterior and posterior hippocampus. Processing was performed using Nifti files converted to ITK-SNAP (http://www.itksnap.org/pmwiki/pmwiki.php) workspaces that were uploaded to the cloud utility offered by ASHS (https://sites.google.com/view/ashs-dox/cloud-ashs/cloud-ashs-for-t1-mri). The resulting segmentations were assessed for quality control of the anterior and posterior hippocampal regions. Thirty-two segmentations had excessive inclusion of the ventricle or choroid plexus: these segmentations were fixed manually by erasing the erroneously labeled voxels in each coronal plane where they appeared. Estimates of volumes were generated from the segmentation files by adding the anterior and posterior hippocampus for the left and right hemispheres.
