## Supplemental Table 1 for "Dementia is strongly associated with medial temporal atrophy even after accounting for neuropathologies"

**Supplementary Table-1.** *Comparison semi-partial correlation coefficients for hippocampal volumes estimated using different neuroimaging pipelines in NACC data (N=120)*. Bold denotes P<0.05. TIV: total intracranial volume. ADNP: Alzheimer's Disease Neuropathology. HS: Hippocampal Sclerosis. CAA: Cerebral Amyloid Angiopathy. VBM: Voxel-Based Morphometry estimate from Computational Anatomy Toolbox (CAT12). FS: FreeSurfer Hippocampal Subfield Segmentation. ASHS: Automatic Segmentation of Hippocampal Subfields T1w.

| | ***Regions*** | ***Hippocampus*** | | |
|---|---|---|---|---|
| ***Variables*** | | *VBM* | *FS* | *ASHS* |
| ***Demographic*** | *Dementia Status at MRI* | **-0.28** | **-0.25** | **-0.21** |
| | *Age* | **-0.27** | **-0.23** | **-0.15** |
| | *Sex* | 0.05 | 0.05 | 0.08 |
| | *Years Education* | 0.08 | 0.09 | 0.16 |
| | *Years MRI to Death* | 0.12 | 0.07 | 0.06 |
| | *TIV* | **0.22** | **0.30** | **0.26** |
| ***Pathology*** | *ADNP* | -0.06 | -0.07 | -0.03 |
| | *HS* | **-0.32** | **-0.27** | **-0.29** |
| | *Lewy Bodies* | -0.05 | -0.01 | -0.04 |
| | *CAA* | -0.06 | -0.01 | -0.02 |
| | *Atherosclerosis* | -0.06 | -0.02 | -0.06 |
| *Adjusted $R^2$* | | 0.40 | 0.40 | 0.31 |
