## Supplemental Table 2 for "Dementia is strongly associated with medial temporal atrophy even after accounting for neuropathologies"

**Supplementary Table-2.** *Comparison of NACC (N=129) and ADNI (N=49) semi-partial correlation coefficients for ROI volumes across the various demographic and neuropathological measures, with dichotomized Braak stages instead of ADNP included in the model*. Bold denotes P<0.05. TIV: total intracranial volume. HS: Hippocampal Sclerosis. Semi-Part. Corr.: semi-partial correlation coefficient.

| ***Dichotomized by Braak Stage V/VI*** | ***Hippocampus Semi-Part. Corr.*** | | ***Amygdala Semi-Part. Corr.*** | | ***Parahippocampus Semi-Part. Corr.*** | |
|---|---|---|---|---|---|---|
| ***Variables*** | *NACC* | *ADNI* | *NACC* | *ADNI* | *NACC* | *ADNI* |
| *Dementia* | **-0.35** | **-0.37** | **-0.33** | -0.23 | **-0.31** | **-0.36** |
| *Age* | **-0.37** | 0.11 | **-0.32** | 0.12 | **-0.34** | 0.11 |
| *TIV* | **0.28** | 0.21 | **0.37** | 0.15 | **0.33** | 0.05 |
| *Braak V/VI* | -0.04 | -0.17 | -0.08 | -0.25 | -0.08 | **-0.27** |
| *HS* | **-0.28** | **-0.46** | **-0.18** | -0.26 | **-0.17** | **-0.33** |
| *Lewy Bodies* | -0.04 | **-0.26** | **-0.16** | -0.24 | -0.07 | -0.20 |

| ***Dichotomized by Braak Stage III/IV*** | ***Hippocampus Semi-Part. Corr.*** | | ***Amygdala Semi-Part. Corr.*** | | ***Parahippocampus Semi-Part. Corr.*** | |
|---|---|---|---|---|---|---|
| ***Variables*** | *NACC* | *ADNI* | *NACC* | *ADNI* | *NACC* | *ADNI* |
| *Dementia* | **-0.35** | **-0.39** | **-0.34** | -0.26 | **-0.33** | **-0.39** |
| *Age* | **-0.37** | 0.13 | **-0.33** | 0.15 | **-0.34** | 0.13 |
| *TIV* | **0.28** | 0.22 | **0.37** | 0.16 | **0.33** | 0.07 |
| *Braak III/IV/V/VI* | -0.06 | -0.10 | -0.07 | -0.21 | -0.03 | -0.20 |
| *HS* | **-0.28** | **-0.47** | **-0.18** | **-0.28** | **-0.17** | **-0.35** |
| *Lewy Bodies* | -0.04 | **-0.27** | **-0.16** | -0.26 | -0.06 | -0.21 |
