## Supplemental Table 3 for "Dementia is strongly associated with medial temporal atrophy even after accounting for neuropathologies"

**Supplementary Table-3.** *Comparison semi-partial correlation coefficients for HS, TDP-43, or both variables in the same model, for ADNI data (N=43)*. Bold denotes P<0.05. TIV: total intracranial volume. ADNP: Alzheimer's Disease Neuropathology. HS: Hippocampal Sclerosis. Semi-Part. Corr.: semi-partial correlation coefficient. TDP-43: TAR DNA-binding protein 43. Hipp.: Hippocampus.

| ***Regions*** | ***Hippocampus Semi-Part. Corr.*** | | | ***Amygdala Semi-Part. Corr.*** | | | ***Parahippocampus Semi-Part. Corr.*** | | |
|---|---|---|---|---|---|---|---|---|---|
| ***Variables*** | *HS* | *TDP-43* | *Both* | *HS* | *TDP-43* | *Both* | *HS* | *TDP-43* | *Both* |
| *Dementia* | **-0.35** | -0.27 | **-0.32** | -0.21 | -0.16 | -0.19 | **-0.33** | -0.28 | **-0.32** |
| *Age* | 0.08 | -0.01 | 0.07 | 0.18 | 0.13 | 0.17 | 0.15 | 0.09 | 0.15 |
| *TIV* | 0.23 | 0.19 | 0.24 | 0.20 | 0.18 | 0.21 | 0.11 | 0.07 | 0.11 |
| *ADNP* | -0.15 | -0.18 | -0.17 | -0.13 | -0.16 | -0.15 | -0.15 | -0.17 | -0.16 |
| *HS* | **-0.49** | | **-0.41** | -0.29 | | -0.24 | **-0.36** | | **-0.31** |
| *TDP-43 Hipp.* | | **-0.30** | -0.14 | | -0.20 | -0.10 | | -0.20 | -0.08 |
| *Lewy Bodies* | **-0.26** | -0.14 | **-0.26** | -0.28 | -0.22 | -0.28 | -0.23 | -0.14 | -0.23 |
| *Adjusted $R^2$* | **0.33** | 0.15 | **0.33** | 0.12 | 0.06 | 0.10 | **0.21** | 0.11 | **0.20** |
